# Efficacy of Short vs. Long Dog-Assisted Therapy Programs in Children and Adolescents with Fetal Alcohol Spectrum Disorder: A Randomized Controlled Trial

**DOI:** 10.1101/2025.04.26.25326504

**Authors:** Sílvia Muñoz, Raquel Vidal, Jorge Lugo, Francesc Ristol, Anna Veiga, Cristina Vico, Josep Antoni Ramos-Quiroga, Nuria Gómez-Barros

**Affiliations:** Department of Psychiatry, Hospital Universitari Vall d’Hebron, Barcelona, Catalonia. Spain; Group of Psychiatry, Mental Health and Addiction, Vall d’Hebron Research Institute (VHIR); Biomedical Network Research Centre on Mental Health (CIBERSAM), Instituto de Salud Carlos III; Department of Psychiatry and Legal Medicine, Universitat Autònoma de Barcelona; Centre de Terapia Assistida amb Cans (CTAC); Fundació Probitas

## Abstract

The objective of this study was to compare the efficacy of a shorter (8 sessions) and longer (16 sessions) Dog-Assisted Therapy (DAT) program for children and adolescents with Fetal Alcohol Spectrum Disorder (FASD). We evaluated the impact of DAT on social skills, internalizing and externalizing problems, quality of life, severity of the disorder, and parental emotional well-being. A randomized controlled trial was conducted with 55 FASD patients, assigned to either the short (n = 24) or long (n = 31) DAT program. The longer DAT group showed significant reductions in externalizing symptoms (CBCL inattention; F = 4.676; p = 0.035; ES = 0.083) and greater improvements in social skills (SSIS-P Problem Behavior: F = 7.803, p = 0.007, ES = 0.13), lower FASD severity scores (CGI-S Clinician; F = 6.54, p = 0.014, ES = 0.014; CGI-S Parents: F = 4.938, p = 0.031; ES = 0.087), and enhanced quality of life (K-Screen Peers and Social: F = 4.38, p = 0.04, ES = 0.78) compared to the shorter program. Parents in the long DAT group also reported significant reductions in depressive symptoms (BDI-II; F = 14.03, p = 0.000, ES = 2.12). These findings suggest that both short- and long-duration DAT programs are effective, with longer programs offering additional benefits in specific domains.

**Clinical Trial Registration:** NCT06763614.

## INTRODUCTION

Fetal Alcohol Spectrum Disorder (FASD) is a neurodevelopmental condition resulting from prenatal alcohol exposure, characterized by cognitive, emotional, physical, and behavioral impairments. It affects approximately 1% to 5% of the population (May et al., 2018) and is often comorbid with Attention Deficit Hyperactivity Disorder (ADHD) and Autism Spectrum Disorder (ASD) (Singal et al., 2018). The lifelong challenges associated with FASD, including intellectual disability, social skill deficits, and behavioral problems, place significant emotional and caregiving burdens on parents (Mukherjee et al., 2013).

Previous research has highlighted the effectiveness of psychological intervention programs focused on emotional regulation for individuals with FASD (Wells et al., 2012; Wells, 2009). Similarly, studies have emphasized the benefits of interventions targeting social skills (Keil et al., 2010; O’Connor et al., 2006). These interventions have been shown to result in reductions in externalizing behaviors, with social skills training specifically correlating with improvements in social competence and a decrease in problematic behaviors Animal-Assisted Therapy (AAT), particularly Dog-Assisted Therapy (DAT), has gained recognition for its potential to improve psychosocial functioning in individuals with neurodevelopmental disorders (Narvekar & Krishnan, 2024; Narvekar & Narvekar, 2022). DAT leverages the human-animal bond to enhance emotional regulation, social skills, and overall well-being (Adams, 2009).

The association between delivering AATs to patients with neurodevelopmental disorders and positive outcomes has been acknowledged in the literature (Carlisle et al., 2025; Gómez-Calcerrada et al., 2021; Schuck et al., 2018). Moreover, studies have recently demonstrated the efficacy of DAT in reducing externalizing symptoms and improving social skills in children with FASD (Vidal et al., 2023; Vidal et al., 2020). However, the optimal duration of DAT programs remains unclear.

This study aims to compare the efficacy of a short (8 sessions) and long (16 sessions) DAT program for children and adolescents with FASD. We hypothesize that the longer DAT program will yield superior outcomes in reducing externalizing symptoms, improving social skills, and enhancing quality of life. Additionally, we anticipate greater reductions in parental anxiety and depressive symptoms with the longer treatment.

## MATERIALS AND METHODS

### Study Design

The design was a randomized, rater-blinded, controlled trial. Participants were randomly assigned either to the long version group of 16 sessions (n = 31) or to the short version group of 8 sessions of DAT treatment (n = 24). A random allocation sequence was generated using computer-generated random numbers, ensuring equal chances for participants to be assigned to either short or long treatment. Block randomization ensured balanced allocation based on key demographic variables, such as age, to minimize potential confounding factors. To prevent selection bias, allocation concealment was implemented, ensuring that those enrolling participants were unaware of group assignments until allocation. Raters assessing outcomes were blinded to the participants’ group assignments.

### Participants

The study recruited patients from the university hospital’s FASD Program. The sample size was determined based on the number of patients available and willing to participate within the study’s timeframe (Jan 2024 to Jan 2025). We included patients with FASD diagnosis, between 6 and 18 years of age, with or without other comorbid psychiatric disorders. As inclusion criteria, all participants needed to have stable doses of medication for at least two months before the study and agree not to seek out any other psychiatric or psychological treatment during the study. Patients exhibiting behavioral instability, aggressive tendencies, or requiring intensive treatment (hospitalization or day hospital) were excluded. Considering the cognitive challenges associated to FASD, we found it pertinent to involve also individuals with borderline IQ or intellectual disabilities.

Participants whose behavior may result in aggressive or inappropriate interactions with dogs, posing risks to both the child and the animal were excluded. Children and teenagers with fear of dogs, which could compromise their safety and comfort; allergies to dogs or other animals, which could lead to adverse reactions; behavioral or psychological conditions medical conditions that could be exacerbated by the presence of dogs or interfere with the study’s objectives; and inappropriate behaviors towards animals, such as hitting or teasing dogs, which would not ensure positive and safe interactions could not participate.

### Intervention

Short and long DAT program comprised 8 and 16 manualized sessions respectively. Both versions were divided into two phases: individual and group intervention. CTAC Method (Center of Dog Assisted Therapy) was implemented. CTAC is a full-member of the International Association of Human-Animal Interaction Organizations (IAHAIO), a global association of organizations that engage in practice, research and/or education in dog assisted therapy. Interventions were administered weekly and each lasted 45 minutes.

### Treatment Content

Following the initial encouragement of interaction between the patient and the therapy dog to foster companionship and trust, the individual phase focused on skill development, covering cognitive abilities such as organization and planning (sequential thinking), attention and concentration, emotional regulation, impulsivity management, problem-solving, and content review. The group-based sessions entailed organized interventions concentrating on social skill elements, including empathy, assertiveness training, adaptive behavior, handling criticism, communication, and teamwork.

Both DAT versions included two phases—individual and group interventions—with the longer version offering extended, content-related sessions. The individual modules were structured with the following session sequence and contents: S1 focused on building and establishing a bond patient-animal; S2 covered executive functions; S3 addressed organization and planning skills; S4 targeted frustration tolerance; S5 addressed emotional self-regulation; S6 focused on managing impulsivity; S7 covered problem-solving techniques; and S8 reviewed the material covered. Following the individual interventions, groups of 3 to 5 patients were inmediately allocated. The eight group sessions focused on social skills: S9 introduction on social skills, S10 assertiveness training, S11 criticism management, S12 Cooperation skills, S13 Empathy, s14-15 Social Skills training and S16 content review.

The sessions involved the presence of two certified therapy dogs, alongside a specialized technician in DAT, and a psychologist who acted as a facilitator in both the individual and the group modules. Patients received visits from their psychiatrist to monitor their adherence to medication.

### Diagnostic and Outcome Measures

An initial evaluation was conducted by a team of experts from various fields, including a geneticist specializing in dysmorphology assessment, a neuropsychologist for cognitive testing, and a psychiatrist for evaluating behavior and any comorbid conditions. All outcome measures were administered both before treatment (T1 baseline) and at the conclusion of treatment (T2).

The variables of the present study included primary outcomes, which were assessed to measure the main objectives:

- Internalizing and Externalizing symptoms: the Parent Version of the Child Behavior Checklist (CBCL) (Achenbach & Edelbrock, 1991) assessed internalizing symptoms (e.g., anxiety/depression) and externalizing symptoms (e.g., impulsivity, oppositional behaviors). This 113-item scale evaluates withdrawn (e.g., behaving too immaturely, uncooperative tendencies), somatic complaints (e.g., stomachaches, headaches), anxiety/depressive symptoms (e.g., feelings of being hurt, upset, or nervous), thought-related issues (e.g., distracted mind, repeating actions), ADHD-related traits (e.g., impulsivity, immature behavior), oppositional behaviors (e.g., defiance, arguing), and general behavioral challenges (e.g., lack of guilt, vandalism).
- Social Skills: Social Skills Improvement System-Parent Form (SSIS-P) (Gresham & Elliott, 2008) was used to evaluate this variable. It is a scale consisting of 79 items evaluates social skills and problematic behaviors in children and adolescents as reported by their parents. Within the Social Skills domain, subscales encompass communication, cooperation, assertion, responsibility, empathy, engagement, and self-control (e.g., expressing feelings when wronged, seeking help from adults, interacting positively with peers, attempting to console others). The problematic behaviors domain includes both internalizing and externalizing issues, bullying, hyperactivity/inattention, and autism spectrum traits
- Quality of life: KidScreen 27 (Ravens-Sieberer et al., 2007) Parent version is a comprehensive assessment of children and adolescents’ overall health and quality of life integrating five aspects: Physical Well-Being (involving physical activities and health), Psychological Well-Being (encompassing mood and emotions), Autonomy and Family (related to family and leisure time), Social Interaction with Peers (regarding friendships), and Support within School Environment (involving aspects of education and school life).
- FASD symptoms severity: we used the the Clinical Global Impression Scale for Severity (CGI-S) (Rapoport et al., 1985). It is a 7-point scale (1 = normal, 2 = borderline mentally ill, 3 = mildly ill, 4 = moderately ill, 5 = markedly ill, 6 = severally ill, and 7 = extremely ill) (NIMH, 1985).
- State-Trait Anxiety Inventory (STAI) is a commonly used measure of trait and state anxiety (Spielberger et al., 1971). It can be used in clinical settings to diagnose anxiety and to distinguish it from depressive syndromes.
- The Beck Depression Inventory-II (BDI-II) is a widely used self-report instrument for assessing the severity of depressive symptoms in adolescents and adults aged 13 and over (Beck et al., 1996).

Secondary outcomes were:

- Sociodemographic characteristics: patient’s characteristics (age and gender) were assessed in the clinical interview during the diagnostic evaluation.
- FASD subtype: We used the Hoyme criteria that distinct three FASD subtypes: Complete FAS, pFAS, and ARND (Hoyme et al., 2016).
- Pharmacological treatment: The pharmacological treatment involved documenting and overseeing medication usage to evaluate the patient’s adherence to and ongoing use of medications as prescribed by the psychiatrist.
- Comorbidity: to assess comorbidity we used the semistructured K-SADS interview for children and adolescents aged under 16 (Matuschek et al., 2016) and the Structured Clinical Interview for DSM-IV Axis I and Axis II Disorders (SCID-I and SCID-II) for the evaluation of older adolescents (First & Gibbon, 2004).
- Intellectual Quotient: The Wechsler Intelligence Scale WISC-V or WAIS-V (Wechsler, 1981) according to the patient’s age,was used in order to evaluate global cognitive capacity.

The completion of all measures was done by parents as patients with FASD often exhibit limited self-awareness, and the reliability of their self-reports may be uncertain.

### Procedure

The ethics committee of clinical investigation at the Hospital Universitari Vall d’Hebron approved the study before participant enrollment. The study was conducted between Jan 2024 and Jan 2025. During this period, of 72 patients approached through the FASD unit, 64 consented to participate after receiving detailed information about the study, three declined to participate and five did not meet inclusion criteria. Of the 64 participants enrolled, 55 completed treatment. Written informed consent was obtained from parents and informed assent was obtained from participants/patients. Following the pre-treatment assessment, the study’s data manager utilized a computerized random number generator (SPSS version 20) to randomly assign participants to the two treatment variations. The raters of the study were blinded to the intervention and had no engagement in the trial other than interviewing the participants during the pre-treatment and post-treatment evaluations. Participants in the two groups were evaluated at the beginning of the study (baseline) and at the end of the treatment (post). Pre-test assessment was administered 1 week before the beginning of the intervention and the post-test assessment one week after the intervention that lasted either 8 or 16 weeks.

### Statistical Analysis

Data were analyzed (using SPSS version 20). An analysis of variance for repeated measures was performed, analyzing group and time effect and its interaction effect. The analysis focused on comparing the means of two distinct groups at a single point in time, assuming that the data within each group follows a normal distribution and that the variances between the groups are approximately equal. Additionally, significance tests (t-test and chi-square) were employed to compare baseline characteristics between two arms. All reported results were significant at the 5% level. Partial eta squared (η2) was calculated to estimate the effect size of treatment outcomes, where η2 = 0.01 indicates a small effect. η2 = 0.06 indicates a medium effect. η2 = 0.14 indicates a large effect.

## RESULTS

### Sample Characteristics

The demographic characteristics of participants across the two treatment groups are summarized in Table 1. No statistically significant differences between groups were detected with respect to demographic (gender, age), clinical (FASD subtype, psychiatric comorbidity, IQ,) and baseline measures of the participants.

**Table 1.** Participants’ characteristics.

|  | **Short version**<br>**(n=24)** |  | **Long version**<br>**(n=31)** |  |  |  |
| --- | --- | --- | --- | --- | --- | --- |
| **Variables** | **n** | **%** | **N** | **%** | **p** | **X<sup>2</sup>** |
| **Gender** |  |  |  |  | 0.732 | 0.117 |
| Male | 14 | 58.3 | 16 | 51.61 |  |  |
| Female | 10 | 41.66 | 15 | 48.38 |  |  |
| **Comorbidity** |  |  |  |  |  |  |
| ADHD | 22 | 91.67 | 29 | 93.5 | 0.79 | 0.071 |
| ASD | 3 | 12.5 | 6 | 19.35 | 0.49 | 0.46 |
| Learning Disorder | 7 | 29.17 | 5 | 16.13 | 0.25 | 1.35 |
| **Diagnosis** |  |  |  |  |  |  |
| Complete FASD | 9 | 37.5 | 19 | 61.3 | 0.08 | 3.063 |
| Partial FASD | 11 | 45.83 | 9 | 29.0 | 0.2 | 1.65 |
| ARND | 4 | 16.67 | 3 | 9.7 | 0.44 | 0.595 |
|  | **Mean** | **SD** | **Mean** | **SD** | **p** | **t** |
| **Age** | 14.33 | 1.94 | 13.06 | 2.75 | 0.61 | 1.91 |
| **IQ** | 75.25 | 14.83 | 72.16 | 11.076 | 0.380 | .885 |

*FAS, Fetal Alcohol Syndrome; ARND, Alcohol Related Neurodevelopmental Disorder; ADHD, Attention Deficit Hyperactivity Disorder; ASD, Autism Spectrum Disorder; IQ, Intellectual Quotient*

In terms of gender distribution, the short treatment group comprised 58.3 % males and 41.6 % females, while the long treatment group had 51.6 % males and 48.3% females. The chi-square test revealed no significant difference in gender distribution between the groups (χ^2^(1) = 0.117, p = 0.732), indicating a comparable representation of genders in both treatment conditions.

Participant comorbidity profiles were examined to understand the prevalence of various conditions within each treatment group. Notable comorbidities included Attention-Deficit/Hyperactivity Disorder (ADHD) (Short: 91.67%, Long: 93.5%), autism spectrum disorder (ASD) (Short: 12.5%, Long: 19.35%), and learning disorders (Short: 29.17%, Long: 16.13%). The chi-square test demonstrated no significant difference in comorbidity distribution between the two treatment groups.

Diagnoses were further explored, in the short treatment group, 45.83% of participants received a diagnosis of partial FAS, 37.5% complete FAS, and 16.67% Alcohol-Related Neurodevelopmental Disorder. Conversely, the long treatment group exhibited a distribution of 29.0% partial FAS, 61.3% complete FAS, and 9.7% ARND. The chi-square test demonstrated no significant difference in diagnosis distribution between the two treatment groups.

Analysis of age revealed a mean age of 14.33 years (SD = 1.94) for the short treatment group and 13.06 years (SD = 2.75) for the long treatment group. The independent samples t-test indicated no statistically significant difference neither in age or IQ.

### Program Completion Rate

A flow diagram of the study is shown in Figure 1. Low dropout rates were observed for the two groups.

**Figure 1.**
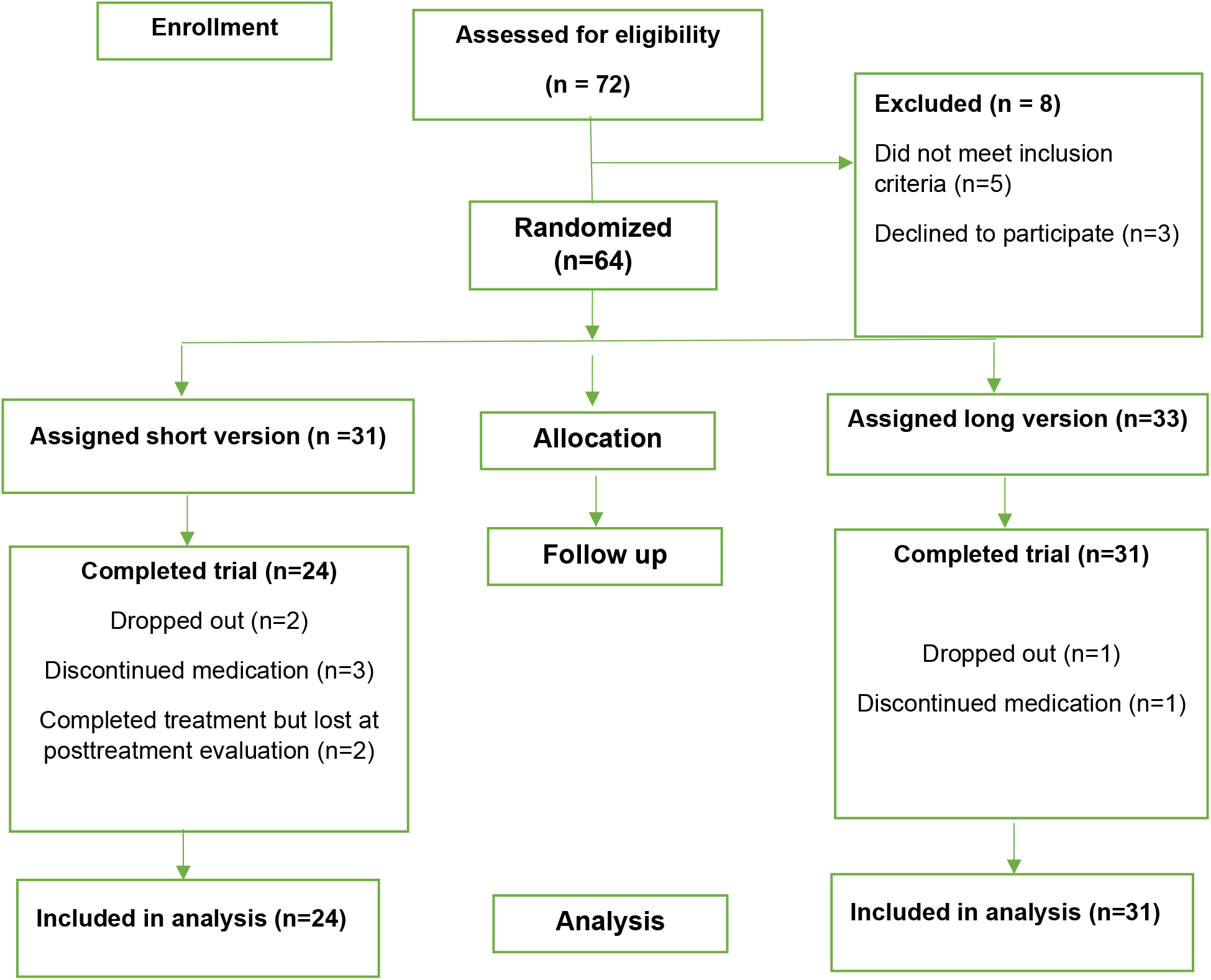
Flow diagram through the phases of the study.

### Outcome Measures

Table 2 represents the baseline and outcome measures for participants in the short version (n = 24) and the long version (n = 31) groups. Primary outcomes of the study were:

**Table 2.** Outcome results.

| Measures | Short version(n = 24) |  | Longversion (n = 31) |  |  |  |  |  |
| --- | --- | --- | --- | --- | --- | --- | --- | --- |
|  | Baseline | Outcome | Baseline | Outcome | Time |  | Time x Group |  |
|  | Mean (SD) | Mean (SD) | Mean (SD) | Mean (SD) | *F(p)* | *ES* | *F(p)* | *ES* |
| **CBCL Internalizing** | 66.79 (8.01) | 65 (7.49) | 64.47 (7.92) | 61.87 (6.43) | 7.279 (.009)** | .123 | .247 (.622) | .005 |
| Withdrawn | 67.83 (12.08) | 62.75 (10.26) | 64.37 (10.76) | 60.4 (7) | 13.22 (.001)* | .203 | .201 (.656) | .004 |
| Somatic Complaints | 52 (2.28) | 51.54 (2.32) | 53.40 (4.72) | 53.43 (5.13) | .322 (.573) | .006 | .431 (.514) | .008 |
| Anxiety/Depression | 68.79 (8.9) | 67.71 (9.17) | 65.67 (7.93) | 63.40 (7.58) | 3.911 (.053) | .070 | .488 (.488) | .009 |
| Social Problems | 73 (9.3) | 69.67 (9.18) | 76.10 (8.23) | 72.47 (9.41) | 16.29 (.000)** | .242 | .004 (.950) | .000 |
| Thought Problems | 67 (7.98) | 65.38 (7.79) | 63.20 (7.68) | 62.03 (7.9) | 1.567 (.216) | .029 | .042 (.838) | .001 |
| **CBCL Externalizing** | 66.63 (7.10) | 64.79 (6.81) | 64.33 (10.43) | 61.60 (9.93) | 13.178 (.001)* | .202 | .512 (.478) | .010 |
| Inattention | 73.08 (8.35) | 73.42 (6.68) | 73.27 (7.52) | 69.73 (7.55) | 3.203 (.079) | .079 | 4.676 (.035)* | .083 |
| Opposition | 65.46 (9.76) | 63.83 (10.22) | 67.03 (11.73) | 64 (10.20) | 12.128 (.001)* | .189 | 1.108 (.297) | .021 |
| Conduct Problems | 63.95 (8.33) | 60.87 (7.86) | 62.8 (7) | 59.23 (6.63) | 34.559 (0.000)* | .399 | 0.183 (0.671) | .003 |
| **CBCL total** | 70.17 (3.85) | 67.88 (4.46) | 67.47 (7.24) | 64.50 (7.73) | 19.847 (.000)** | .276 | .327 (.570) | .006 |
| **SSIS-P Social Skills** | 69.88 (15.58) | 73.67 (15.75) | 65.63 (16.37) | 73.33 (18.32) | 21.009 (.000)** | .288 | 2.430 (.125) | .045 |
| **SSIS-P Problem Behavior** | 42.33 (10.32) | 39.17 (10.22) | 47.80 (16.04) | 39.63 (12.57) | 40.088 (.000)** | .435 | 7.803 (.007)** | .130 |
| **KID SCREEN Total** | 70.12 (9.32) | 67 (8.41) | 69 (9.12) | 66 (9.21) | 9.729 (.003)** | .158 | .144 (.706) | .003 |
| Physical well-being | 12.08 (4.47) | 11.33 (3.96) | 13.70 (3.46) | 12.93 (4.49) | 3.38 (.072) | .061 | .000 (.984) | .000 |
| Psychological | 17.58 (2.96) | 18 (3.2) | 17 (4.67) | 17.57<br>(4.03) | .967 (.330) | .018 | .023 (.881) | .000 |
| Autonomy and Parents | 20.08 (4.58) | 19.42 (4.34) | 19.23 (4.09) | 18.43<br>(3.60) | .226 (.142) | .041 | .018 (.893) | .000 |
| Peers and Social | 9.67 (2.58) | 9.46 (2.06) | 9.63 (2.67) | 8.33 (2.94) | 8.379<br>*(.006)* | .139 | 4.389<br>(.041)* | .078 |
| Support and School | 9.79 (2.77) | 9.46 (2.9) | 9.13 (2.73) | 9.4 (2.95) | .025 (.876) | .000 | 1.994<br>(.164) | .037 |
| **CGI – S – Clinician** | 4.13 (0.95) | 3.11 (0.71) | 4.20 (0.76) | 3.31 (0.72) | 55.877<br>(.000)** | .518 | 6.54<br>(.014)* | .112 |
| **CGI – S- Parent** | 4.12 (0.88) | 3.13 (0.81) | 3.73 (0.87) | 2.72 (0.75) | 54.023<br>(.000)* | .510 | 4.938<br>(.031)* | .087 |
| **BDI – II Parents** | 19.75 (9.72) | 19.02 (10.04) | 18.63 (9.19) | 15.57<br>(7.37) | 6.137<br>(.017)* | .106 | 14.030<br>(.000)** | .212 |
| **STAI. State anxiety.**<br>**Parents** | 24.29 (11.18) | 21.75 (10.03) | 26.47 (9.55) | 22 (8.56) | 40.091<br>(.000)** | .435 | 3.025<br>(.088) | .055 |
| **STAI. Trait anxiety.**<br>**Parents** | 23.25 (8.87) | 21.79 (7.95) | 24.23 (9.65) | 22.03<br>(10.87) | 12.070<br>(.001)** | .188 | .496 (.484) | .009 |

*CBCL, Child Behavior Checklist; K – Screen, Kids Screen test; SEHS, Social and Emotional Health Survey; CGI – S, Clinical Global Impression Scale, p, significance on time-group Effect, BDI, Beck Depression Inventory; STAI, The State – Trait Anxiety Inventory, \*p < .05; \*\*p < .01. ES, Effect size, partial eta squared effect size where 0.01= small, 0.06= medium, 0.14= large;*

### Internalizing and Externalizing Symptoms

Patients assigned to the long version of DAT group experienced significant reduction in externalizing symptoms (CBCL Inattention; F(1,54) = 4.676; p = 0.035; ES = 0.083) as compared to the short version in the time x group effect analysis. However, the two groups obtained statistically significant differences in the time effect analysis (differences between baseline and prostreatment evaluation regardless group condition): (CBCL Internalizing: F= 7.279, p=0.009, ES=0.123; CBCL withdrawn: F=13.22, P=0.001, ES=0.203; CBCL Social Problems: F=16.29, p= 0.000, ES=0.242; CBCL Externalizing: F=13.17, p=0.001, ES=0.202; CBCL Opposition: F=12.128, p=0.001, ES=0.189).

### Social skills

Patients assigned to the long version of DAT group showed significant improvement on social skills (SSIS-P Problem Behavior; F=7.803, p=0.007, ES= 0.13) as compared to the short version. However, the two groups improved in the time effect analysis on SSIS –P Social Skills (F=21.009, p=0.000, ES= 0.288) ans SSIS-P Problem Behaviour (F= 40.08, p=0.000, ES=0.435).

### Severity of FASD

Patients assigned to the long version of DAT group showed lower scores on FASD severity (CGI-S Clinician; F=6.54, p=0.014, ES=0.014 and CGI-S Parents; F=4.938, p= 0.031; ES= 0.087) as compared to the short version. However, the time effect analysis revealed that patients assigned to the short group also improved in this domain (CGI-S Clinician; F=55.87, p=0.000, ES=0.5 and CGI-S Parents: F=54.023, p=0.000, ES=0.51).

### Quality of life

Patients assigned to the long version of DAT group showed improvements on quality of life (K-Screen Peers and Social; F=4.38, p=0.04, ES=0.78) as compared to the short version of treatment. However, the time effect analysis revealed that patients assigned to the short group also improved in this domain (K-Screen Peers and Social; F=8.379, p=0.006, ES=0.139). The two groups showed improvements on quality of life (KScreen Total: F=9.729, p=0.003, ES=0.158).

### Parent’s anxiety and depression

Parents of patients assigned to the long version showed a significant decrease in depressive symptoms compared to the short version (BDI-II: (BDI-II, F=14.03, p=0.000, ES= 2.12). Although no differences were observed on anxiety symptoms between groups, the two groups showed significant decreases in the time effect analysis (STAI State: F=40.091, p=0.000, ES=0.435 and STAI Trait=F=12.07, p=0.001, ES=0188).

No serious adverse events (SAEs) or harms were reported during the study. All participants tolerated the Dog-Assisted Therapy (DAT) sessions well, and no incidents involving the therapy dogs or participants occurred. Both the short and long treatment groups completed their respective sessions without any significant issues or negative outcomes related to the intervention.

## DISCUSSION

The current study aimed to investigate the differential impact of a short and a long version of DAT program on children and adolescents with FASD using a randomized controlled trial design. The goal was to discern not only if DAT is beneficial but also to identify which duration had the most significant impact and to understand the specific aspects that showed improvement.

Initially, we hypothesized that the long version of the treatment would show better results as compared to the short version. Our first hyphothesis was partially supported because the findings showed that both durations yielded positive outcomes, indicating that DAT, regardless of duration, can be a beneficial therapeutic approach for patients with FASD. The examination of treatment effects allowed us to discern that, while both durations were effective, the 16-week long version showed some distinctive advantages in certain outcome measures but not at all domains.

The long version showed more improvements on inattention problems, on social skills, quality of life and FASD severity than the short version. Thus, we could hyphothethise that these domains should require more sessions to achieve more improvements. Similarly, our previous research on DAT showed improvements on inattention, on social skills, obtained lower scores on FASD severity (Vidal et al. 2020) and also improvements on quality of life (Vidal et al. 2023). The DAT program of our previous research include 12 and 16 sessions respectively (long versions) and the results on externalizing, internalizing symptoms and social skills were along the same lines. This could be explained by the fact that maybe these domains require a more extensive program than other externalizing symptoms such as opposition, conduct problems, withdrawn, etc. On the other hand, patients in the two groups showed improvements on quality of life regardless their treatment condition. This suggests that DAT not only addresses specific behavioral and emotional challenges, it can contribute to a more global improvement in the child’s clinical profile.

Our second hyphotesis predicted that parents would get more benefit from the long treatment version in terms of lessening their anxiety and depressive symptoms. We obtained more improvements on depressive symptoms but not on anxiety. This suggests that in order to diminish anxiety symptoms the two versions could be effective; however, more sessions are needed to improve mood symptoms. Nevertheless, the current reserach added that DAT can also improve parents’ wellbeing, differently from previous studies on DAT for FASD patients that did not include this variable in their design. This suggests that DAT interventions not only impact the child directly but may also contribute to a positive shift in the overall family environment.

While this study provides valuable insights, it is essential to acknowledge certain limitations. The sample size may influence the generalizability of findings. Secondly, we included patients with pharmacological treatment, so it is still unknown if DAT could be a suitable treatment for non-medicated patients with FASD. Additionally, the study focused on immediate post-treatment outcomes, therefore longitudinal assessments would enhance our understanding of the manteinance of DAT effects.

Despite these limitations, the outcomes of this study hold significant implications for clinical practice. The positive effects observed across multiple outcome measures highlight the potential of DAT as a tailored and effective intervention for children and adolescents with FASD. Future research should further explore the optimal duration and intensity of DAT interventions, considering individual differences and potential moderators of treatment response

To conclude, this study contributes to the growing body of evidence supporting the efficacy of DAT in addressing the complex challenges associated with FASD. The positive outcomes on child-level measures, coupled with improvements in parental well-being, underscore the potential of DAT as an holistic therapeutic approach. Integrating DAT into comprehensive treatment plans for children and adolescents with FASD warrants further exploration, with an emphasis on tailoring interventions to individual needs and conducting longitudinal assessments to ascertain long-term benefits.

## Data Availability

Data cannot be shared publicly because it contains sensitive personal and health-related information about minors. Data are available from the Vall d'Hebron Institutional Data Access / Ethics Committee (contact via) for researchers who meet the criteria for access to confidential data.

https://vhir.vallhebron.com/ca

